# Evaluating the frequency and the impact of pharmacogenetic alleles in an ancestrally diverse Biobank population

**DOI:** 10.1101/2022.08.26.22279261

**Authors:** Shefali S. Verma, Karl Keat, Binglan Li, Glenda Hoffecker, Marjorie Risman, Regeneron Genetics Center, Katrin Sangkuhl, Michelle Whirl-Carrillo, Scott Dudek, Anurag Verma, Teri E. Klein, Marylyn D. Ritchie, Sony Tuteja

## Abstract

**Background:** Pharmacogenomics (PGx) aims to utilize a patient’s genetic data to enable safer and more effective prescribing of medications. The Clinical Pharmacogenetics Implementation Consortium (CPIC) provides guidelines with strong evidence for 24 genes that affect 72 medications. Despite strong evidence linking PGx alleles to drug response, there is a large gap in the implementation and return of actionable pharmacogenetic findings to patients in standard clinical practice. In this study, we evaluated opportunities for genetically guided medication prescribing in a diverse health system and determined the frequencies of actionable PGx alleles in an ancestrally diverse biobank population.

**Methods:** A retrospective analysis of the Penn Medicine electronic health records (EHRs), which includes ∼3.3 million patients between 2012-2020, provides a snapshot of the trends in prescriptions for drugs with genotype-based prescribing guidelines (‘CPIC level A or B’) in the Penn Medicine health system. The Penn Medicine BioBank (PMBB) consists of a diverse group of 43,359 participants whose EHRs are linked to genome-wide SNP array and whole exome sequencing (WES) data. We used the Pharmacogenomics Clinical Annotation Tool (PharmCAT), to annotate PGx alleles from PMBB variant call format (VCF) files and identify samples with actionable PGx alleles.

**Results:** We identified ∼316,000 unique patients that were prescribed at least 2 drugs with CPIC Level A or B guidelines. Genetic analysis in PMBB identified that 98.9% of participants carry one or more PGx actionable alleles where treatment modification would be recommended. After linking the genetic data with prescription data from the EHR, 14.2% of participants (n=6157) were prescribed medications that could be impacted by their genotype (as indicated by their PharmCAT report). For example, 856 participants received clopidogrel who carried *CYP2C19* reduced function alleles, placing them at increased risk for major adverse cardiovascular events. When we stratified by genetic ancestry, we found disparities in PGx allele frequencies and clinical burden. Clopidogrel users of Asian ancestry in PMBB had significantly higher rates of *CYP2C19* actionable alleles than European ancestry users of clopidrogrel (p<0.0001, OR=3.68).

**Conclusions:** Clinically actionable PGx alleles are highly prevalent in our health system and many patients were prescribed medications that could be affected by PGx alleles. These results illustrate the potential utility of preemptive genotyping for tailoring of medications and implementation of PGx into routine clinical care.

## Introduction

The vision of precision medicine involves using genomic information to guide preventive measures and tailor treatment for patients. A major component of this vision is pharmacogenomics (PGx), which aims to provide pharmacotherapy guidance for patients carrying genetic alleles impacting response to medications, with the goal of genetically tailored prescribing at the point of care. Many medications, such as opioid analgesics or the anticoagulant warfarin, require careful dose titration to achieve a therapeutic dose, and consideration of PGx information can help select the most appropriate drug at the right dose, avoiding harmful adverse drug reactions. The Clinical Pharmacogenetics Implementation Consortium (CPIC) has published guidelines for 71 drugs based on a large body of evidence showing the impact of PGx alleles which provide specific drug recommendations to alter prescribing practices.(1,2) CPIC also assigns preliminary prioritization levels (called “provisional”) based on cursory review of evidence including, but not limited to, actionability in other professional society guidelines, recommendations from FDA-approved drug labels and assessment of evidence from PharmGKB clinical annotations.(3,4) Level A refers to gene-drug pairs where genetic information ***should be used*** for prescribing decisions and alternative therapies, or dosing are highly likely to be effective and safe.(1) Level B refers to pairs where genetic information ***could be used*** to change prescribing and alternative therapies or dosing are extremely likely to be as effective and as safe as non-genetically based dosing.(1) As of June 2022, there are 96 gene-drug pairs with a CPIC level A or B guideline. Many of the alleles in these genes are common in the population and associated with a large effect size and a clearly identifiable drug response phenotype such as metabolizer status. However, one barrier to PGx implementation is the uncertainty about the true clinical burden of PGx alleles, particularly when accounting for ancestry-specific allele frequencies. For example the *CYP2C19*\*2 no function allele (with rs4244285 as a key SNP) is twice as common in Asian ancestry individuals as those with European ancestry, which will have ethical and legal implications for testing prioritization(5) Understanding the prevalence of PGx alleles and the frequency of prescribing drugs impacted by these alleles in multiple ancestral populations will aid in targeting clinical implementation efforts for drug-gene pairs with the greatest impact.

Electronic Health Records (EHRs) are a digital record of a patient’s diagnosed conditions as well as information about the care they have received. EHRs have also become a robust source of real-world data that when linked to large DNA repositories can validate the association of known PGx alleles with drug response phenotypes but also further enable discovery of novel PGx associations. There have been several successful examples using the EHR to identify novel genetic association with drug side effects.(6–8) The University of Pennsylvania Health System (also known as Penn Medicine) is one of the primary health care providers in Philadelphia and the surrounding region. Penn Medicine consists of over 3.3 million patients whose data are stored in an EHR since 2008. In our study period of 2012-2020, we counted 3,384,922 unique patients. Approximately 150,000 of these patients have enrolled in the Penn Medicine BioBank (PMBB) research program as of July 2022 and thus far 43,359 have been genotyped for research purposes on a genome-wide SNP array and sequenced using whole exome sequencing (WES); all participant genetic data is then linked to their EHR data. Prior studies have shown that >95% of individuals carry one or more alleles that interact with drugs from CPIC Level A guidelines (9–11), but the number of patients that will actually be prescribed the drugs impacted by the alleles they carry is challenging to predict. In consideration of the implementation of preemptive PGx into clinical care, we sought to understand the population of Penn Medicine patients that are impacted by PGx alleles and are prescribed PGx affected medications. Considering this, we evaluated the prescribing patterns in the Penn Medicine EHR for a set of medications that are included in CPIC level A and B guidelines. Actionable PGx alleles are ones for which a medication prescribing change is recommended. Therefore, we also evaluated the frequency of actionable PGx phenotypes, the set of alleles that contribute to a clinically meaningful and observable drug response trait (e.g., drug metabolizer status), and are associated with strong or moderate therapeutic recommendations in the CPIC guidelines, in the genotyped PMBB participants. The large number of ancestrally diverse participants in the PMBB also allowed for ancestry stratified analyses to better understand how drug prescription practices based on an individual’s ancestry may influence the burden of unwanted drug response outcomes.

## Methods

### Dataset

The study was approved by the Institutional Review Board of the University of Pennsylvania and complied with the principles set out in the Declaration of Helsinki. All individuals recruited for the Penn Medicine BioBank (PMBB) are patients of clinical practice sites of the University of Pennsylvania Health System. Appropriate consent was obtained from each participant regarding storage of biological specimens, genetic sequencing/genotyping, and access to all available EHR data. Medication data was extracted using Penn Medicine’s Clinical Data Warehouse, the Penn Data Store (PDS), from January 2012-December 2020. We used the CPIC guidelines available prior to December 2021 to select 49 drugs with prescribing recommendations for our study. For each of the drugs (Table S1), we generated extensive medication libraries that contained every permutation of the medication available in the EHR including by drug name (generic, brand name and combinations products), dose and dosage form (e.g. oral, intravenous) and RxNorm identifiers.(12) Unique patients were counted once per year for each medication to get a unique number of prescriptions per year. Yearly prescribing data were normalized by yearly encounters to the health system to obtain a drug exposure rate and account for the growing number of patients in the health system. Drug exposures and encounters were limited to those which occurred when the patient was 18 years of age or older.

### Genotyping and Whole Exome Sequencing

Individuals were genotyped using the Illumina Global Screening array v.2.0 and minimally processed with PLINK to remove sites with marker call rate <95% and samples with overall call rate <90%. Furthermore, any samples with a discordance between genetically defined sex and EHR reported sex were discarded. Genotyping and whole exome sequencing was performed at the Regeneron Genetics Center using an in-house high throughput, fully automated approach for exome capture. Libraries were generated from 100ng of genomic DNA with a mean fragment length of 200 bp. The libraries were then barcoded and amplified in preparation for multiplexed exome capture and sequencing. Following exome capture, pooled samples were sequenced with 75 bp paired end reads with two 10 bp index reads on the Illumina NovaSeq 6000 platform on S4 flow cells. Samples were then QC filtered using the ‘Goldilocks’ filtering procedure as described previously(13).

### Construction of an Integrated Call Set

Genotyping arrays and exome sequencing in isolation both provide limited coverage of PGx sites across the genome. We took a similar, but simplified approach to McInnes *et al*. to generate an integrated call set combining the array and exome data.(14) Based on prior work indicating that whole exome sequencing has a much higher call accuracy than genotyping arrays(15), the integrated call set favored exome calls over array calls in cases where a site was covered by both. The exome was used as the primary sequence, with sites not covered by the exome filled with available calls from the genotyping array using bcftools version 1.15.1(16). Average coverage was assessed for the array, exome, and integrated call sets by counting the fraction of PharmCAT reference PGx sites that were found in each set.

### PGx Analysis

Annotation of PGx alleles was performed using an in-development version of PharmCAT 2.0(17), a bioinformatics tool to annotate PGx alleles from a VCF file. The version of PharmCAT used for this study interprets alleles included in CPIC guideline genes, using the latest CPIC guidelines published in August 2022. PharmCAT-generated annotations for the 43,359 genotyped individuals were used to identify all PMBB individuals with one or more actionable PGx alleles for the studied drugs. An actionable PGx allele refers to a genotype or diplotype that has a pharmacogenomic phenotype which then has a clinical recommendation for a medication prescribing change based on the genotype/diplotype. PharmCAT provides a report for each individual (patient-participant) containing all drug-gene combinations that have guidelines included in the CPIC database with a few exceptions. The version of PharmCAT at the time of this study did not call *G6PD*, which falls on the X-chromosome, as well as *MT-RNR1*, a mitochondrial gene and also did not call the *HLA* region. While we studied the usage rates of *some* drugs which are affected by *HLA* and *G6PD* genotype/phenotype, we could not study the frequencies of PGx alleles in these genes. CPIC guidelines utilize phenotypes to provide dosing recommendations rather than genotypes. PharmCAT performs these genotypes to phenotype conversions based on allele function and these phenotype frequencies are provided in Table 3. Phenotypes are usually listed as poor metabolizer (PM), intermediate metabolizer (IM), normal metabolizer (NM), rapid metabolizers (RM), and ultrarapid metabolizer (UM). For some complex genes such as *CYP2D6*, genotypes are translated into gene activity scores (AS) to better capture residual enzyme activity for some alleles(2). We calculated the AS for *CYP2C9* and *CYP2D6* based on the CPIC allele functionality tables. Using the PharmCAT calls, we performed an ancestry-stratified analysis to identify patterns of allele and phenotype frequency in different populations. Furthermore, for each drug-gene combination we looked at the rates of patients with actionable PGx alleles (i.e., leading to prescribing change recommendations) for drugs which they were prescribed, stratified by ancestry.

PharmCAT does support inclusion of external calls for *HLA, G6PD*, and *CYP2D6*. Due to the limitations of array and exome data, we were unable to call *CYP2D6* CNV and structural variation with any external tool. PharmCAT does not ordinarily call the *CYP2D6* gene, whose phenotype is reliant on copy-number and structural variation in the gene as well as the *HLA* locus which is difficult to call without specialized assays. However, a “research mode” option in PharmCAT allowed us to call *CYP2D6* from SNPs alone. These calls only feature *CYP2D6* star-alleles that do not define or include structural variation or a gene deletion. We conducted a separate analysis using the *CYP2D6* calls to estimate the prevalence of *CYP2D6* reduced function phenotypes in patients treated with relevant drugs, with the notable caveat that we could not measure effects of gene deletion or duplication.

## Results

### Demographics

There were 11,235,263 unique patient encounters to Penn Medicine from January 1, 2012, to December 31, 2020, across 1,896,012 unique patients. The cohort included 56.1% women and 43.9% men. The median age was 56 years. The EHR race and ethnicity demographics were 59.8% White (56.7% self-reported White and 3.2% self-reported Hispanic White), 23.4% Black (22.4% self-reported Black and 1% self-reported as Hispanic Black), 4.2% Hispanic or Latino, and 4% Asian as shown in Supplementary Figure 1a.

The distribution of genetic ancestry in the PMBB is comprised of 69.2% European ancestry (EUR), 25.7% African ancestry (AFR), 1.6% East Asian ancestry (EAS), 1.3% admixed American ancestry (AMR), 1.3% South Asian ancestry (SAS), and 0.9% unknown ancestry (UNK). 49.7% of PMBB patients are female (Supplementary Figure 1b). Differences in the demographics of PMBB and Penn Medicine can be partly attributed to the distinction between genetic ancestry and self-reported race, as well as the recruitment of most genotyped PMBB patients occurring prior to Penn Medicine’s acquisition of multiple hospitals.

### Drug prescription trends in Penn Medicine EHR

During our study period, 723,115 (21.3%) unique patients receiving care at Penn Medicine had at least 1 record for a medication in our list (see Methods), while 316,235 (9.3%) received 2 or more drugs and 146,885 (4.3%) received 3 or more. The count of medications per patient per year is show in Table 1. The most prescribed medication over the study period was ondansetron, with more than 3 ondansetron prescriptions for every 100 encounters. Ondansetron is an anti-emetic medication used to treat nausea and vomiting. (Table 2).

**Table 1:**
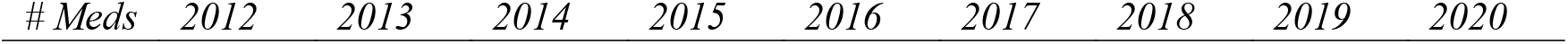
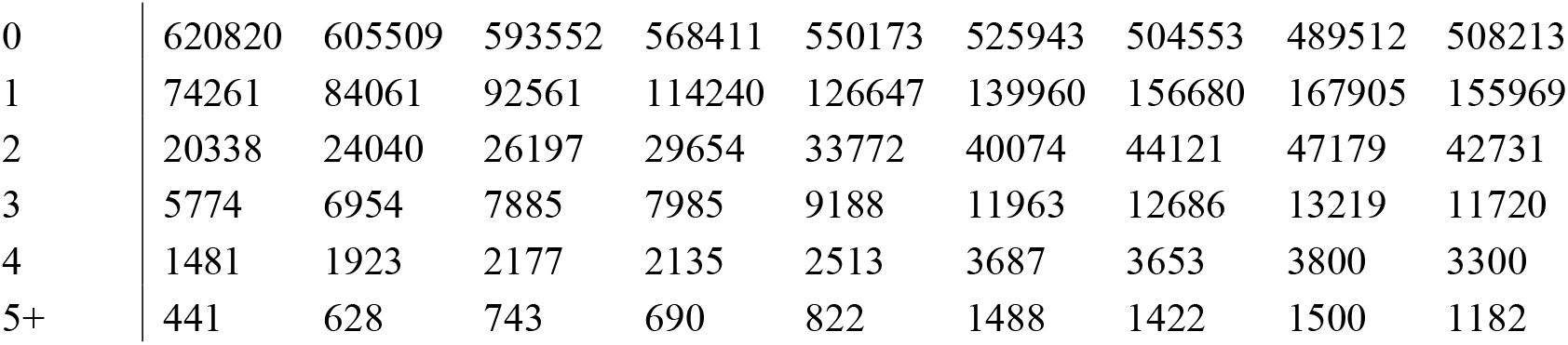
Total number of medications taken by unique patients each year from 2012-2020

**Table 2:**
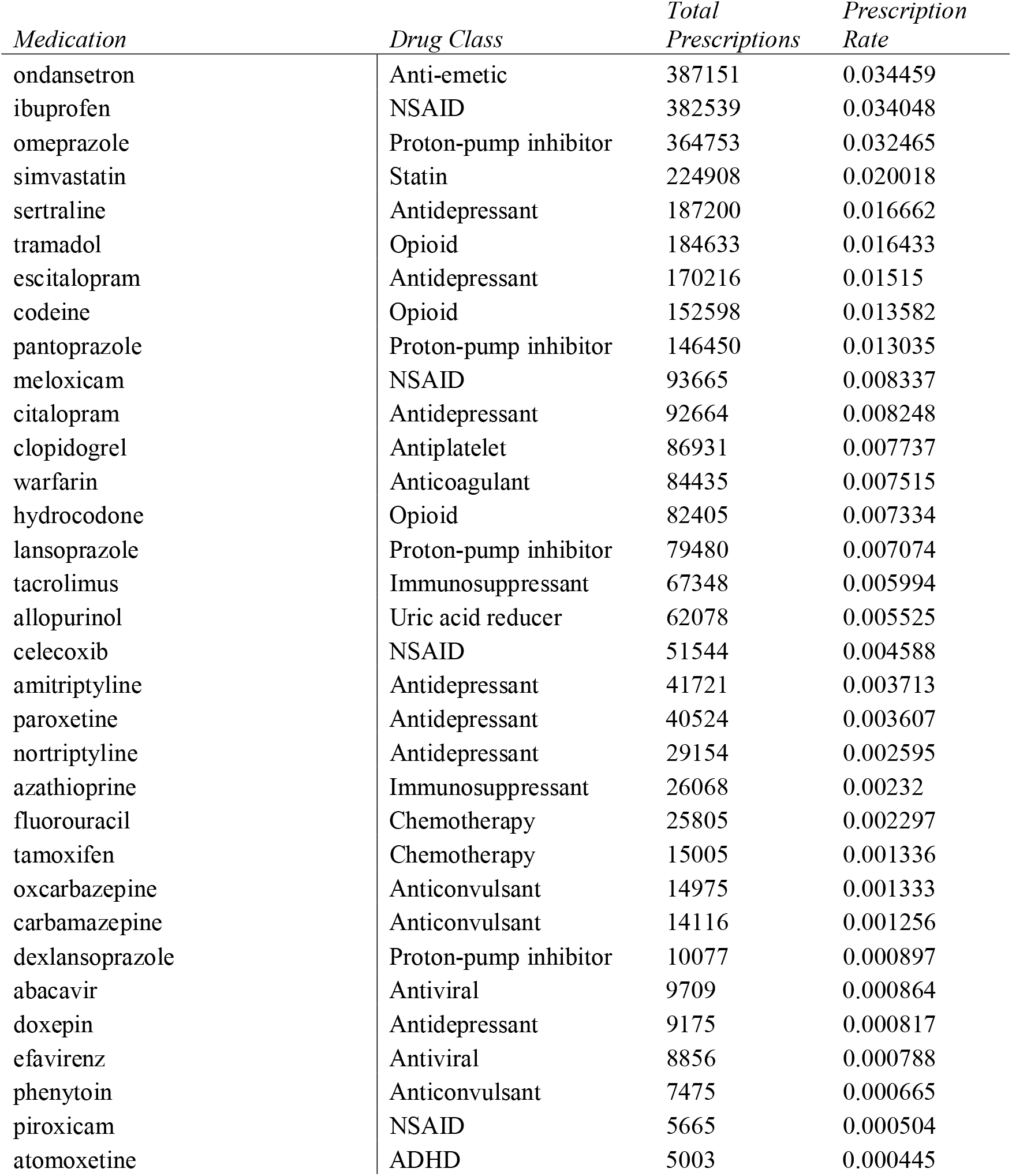
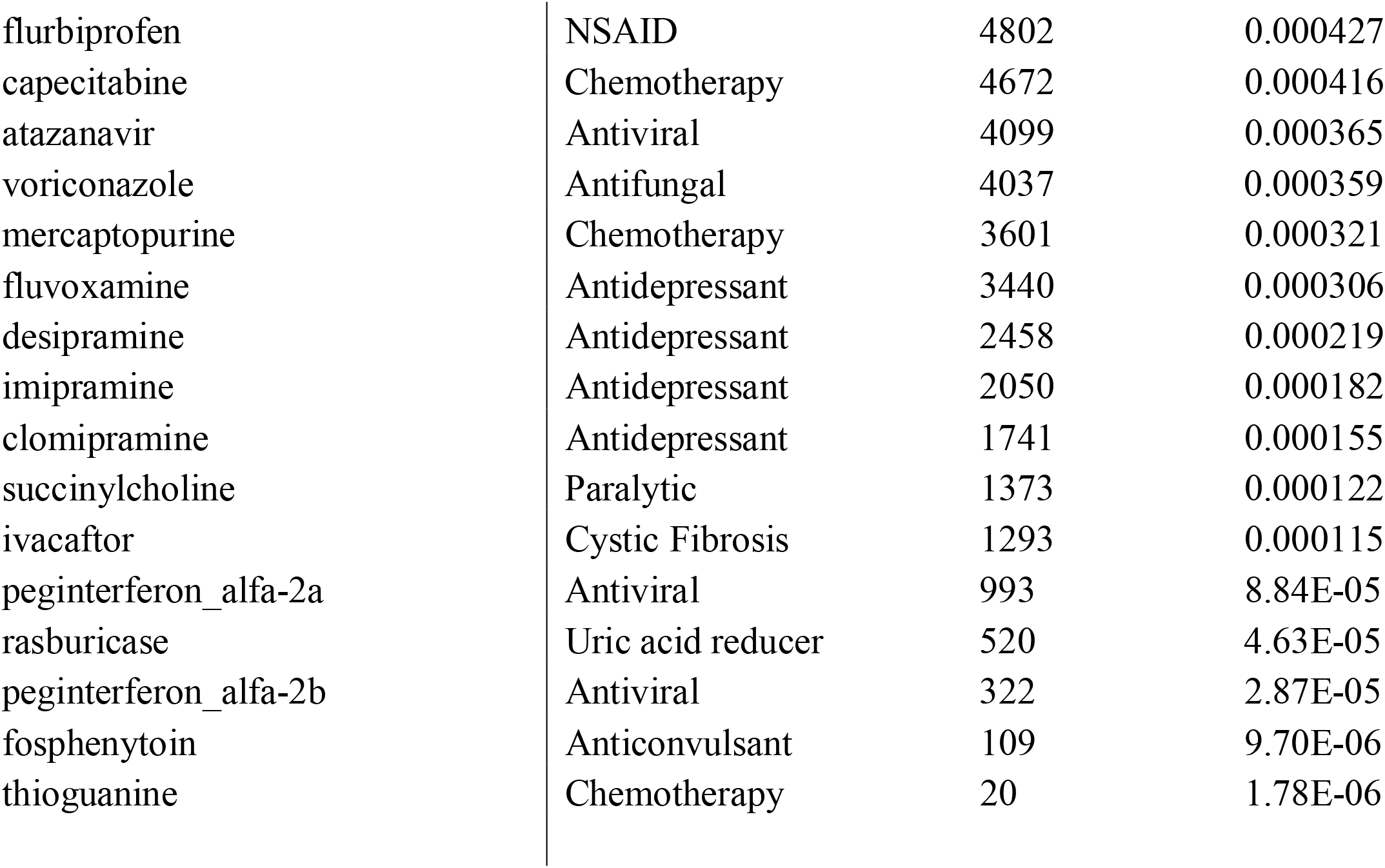
List of PGx informed medications and total prescriptions for each medication. Prescription rate is calculated as total prescriptions/number of unique patient encounters. The list is ordered from high to low prescription rate.

| *Medication* | *Drug Class* | *Total Prescriptions* | *Prescription Rate* |
| --- | --- | --- | --- |
| ondansetron | Anti-emetic | 387151 | 0.034459 |
| ibuprofen | NSAID | 382539 | 0.034048 |
| omeprazole | Proton-pump inhibitor | 364753 | 0.032465 |
| simvastatin | Statin | 224908 | 0.020018 |
| sertraline | Antidepressant | 187200 | 0.016662 |
| tramadol | Opioid | 184633 | 0.016433 |
| escitalopram | Antidepressant | 170216 | 0.01515 |
| codeine | Opioid | 152598 | 0.013582 |
| pantoprazole | Proton-pump inhibitor | 146450 | 0.013035 |
| meloxicam | NSAID | 93665 | 0.008337 |
| citalopram | Antidepressant | 92664 | 0.008248 |
| clopidogrel | Antiplatelet | 86931 | 0.007737 |
| warfarin | Anticoagulant | 84435 | 0.007515 |
| hydrocodone | Opioid | 82405 | 0.007334 |
| lansoprazole | Proton-pump inhibitor | 79480 | 0.007074 |
| tacrolimus | Immunosuppressant | 67348 | 0.005994 |
| allopurinol | Uric acid reducer | 62078 | 0.005525 |
| celecoxib | NSAID | 51544 | 0.004588 |
| amitriptyline | Antidepressant | 41721 | 0.003713 |
| paroxetine | Antidepressant | 40524 | 0.003607 |
| nortriptyline | Antidepressant | 29154 | 0.002595 |
| azathioprine | Immunosuppressant | 26068 | 0.00232 |
| fluorouracil | Chemotherapy | 25805 | 0.002297 |
| tamoxifen | Chemotherapy | 15005 | 0.001336 |
| oxcarbazepine | Anticonvulsant | 14975 | 0.001333 |
| carbamazepine | Anticonvulsant | 14116 | 0.001256 |
| dexlansoprazole | Proton-pump inhibitor | 10077 | 0.000897 |
| abacavir | Antiviral | 9709 | 0.000864 |
| doxepin | Antidepressant | 9175 | 0.000817 |
| efavirenz | Antiviral | 8856 | 0.000788 |
| phenytoin | Anticonvulsant | 7475 | 0.000665 |
| piroxicam | NSAID | 5665 | 0.000504 |
| atomoxetine | ADHD | 5003 | 0.000445 |
| flurbiprofen | NSAID | 4802 | 0.000427 |
| capecitabine | Chemotherapy | 4672 | 0.000416 |
| atazanavir | Antiviral | 4099 | 0.000365 |
| voriconazole | Antifungal | 4037 | 0.000359 |
| mercaptopurine | Chemotherapy | 3601 | 0.000321 |
| fluvoxamine | Antidepressant | 3440 | 0.000306 |
| desipramine | Antidepressant | 2458 | 0.000219 |
| imipramine | Antidepressant | 2050 | 0.000182 |
| clomipramine | Antidepressant | 1741 | 0.000155 |
| succinylcholine | Paralytic | 1373 | 0.000122 |
| ivacaftor | Cystic Fibrosis | 1293 | 0.000115 |
| peginterferon_alfa-2a | Antiviral | 993 | 8.84E-05 |
| rasburicase | Uric acid reducer | 520 | 4.63E-05 |
| peginterferon_alfa-2b | Antiviral | 322 | 2.87E-05 |
| fosphenytoin | Anticonvulsant | 109 | 9.70E-06 |
| thioguanine | Chemotherapy | 20 | 1.78E-06 |

An evaluation of prescribing patterns over time for the studied medications showed an increasing use of some medications, particularly for ondansetron and ibuprofen when the total prescriptions are normalized by the number of encounters for each year as shown in Figure 1. At the same time, despite a large decline in the usage of simvastatin, it remains one of the most prescribed drugs, with more than 1 in 100 encounters involving a prescription. Similarly, while the use of warfarin is declining, it is still used very frequently. Additionally, we examined prescribing patterns for 3 drug classes: non-steroidal anti-inflammatory drugs (NSAIDs), proton-pump inhibitors, and antidepressant drugs (Figure 2).

**Figure 1:**
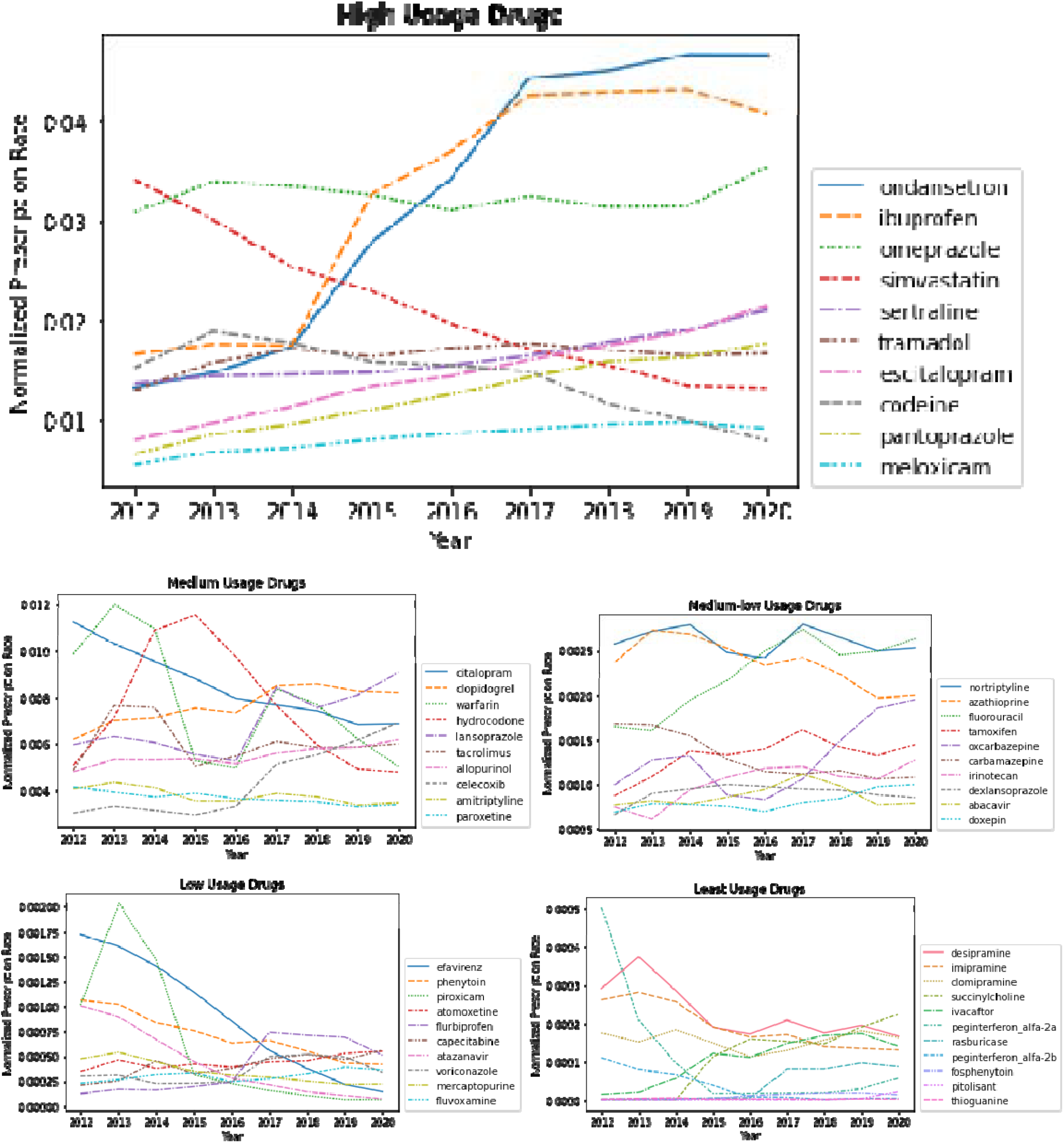
Medication prescription trends over the years of study period. Each line represents a separate medication. Prescription rate is calculated as total prescriptions each year/unique patient encounters each year. The plot is divided into 5 categories based on overall drug usage/prescription rates. The y-axes are scaled differently to reflect the differences in order of magnitude in prescription rates between categories.

**Figure 2:**
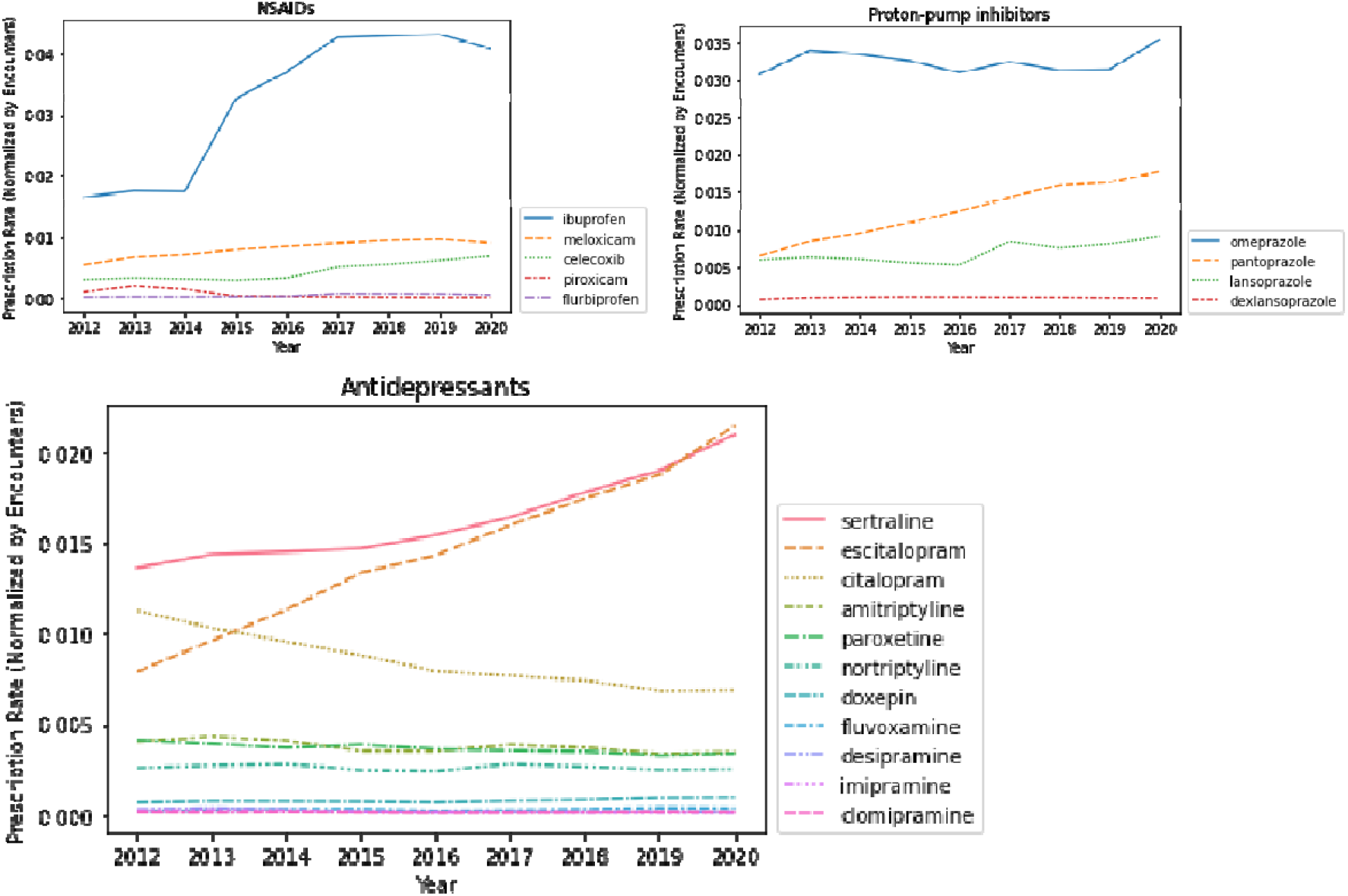
Number of prescriptions for three drug classes (NSAIDs, proton-pump inhibitors, and antidepressants) for each year. Three classes of drugs are in separate panels, x-axis is year and y-axis are the prescription count normalized by the number of encounters in each year. Note that the y-axis scale differs between drug class.

### Integrated call set performance

We compared the performance of our integrated call set to the whole exome sequencing and genotyping array data by measuring their coverage of the PGx sites used by PharmCAT. Of 536 total sites across all samples, the arrays covered an average of 250 sites (46.6%), the exome covered an average of 259 (48.3%) and the integrated call set covered an average of 354 (66%). This validates that exome and genotyping arrays have complementary information for PGx and should be combined, when possible, for larger coverage of the genome. For example, the noncoding variant *rs12248560* is vital to calling the *CYP2C19*17* increased function allele, which increases *CYP2C19* gene expression. In combination with a normal function allele, the *\*17* allele results in a RM phenotype. In case of two *\*17* alleles, there is an UM phenotype. Lastly, combinations of *\*17* with reduced function or loss of function alleles can result in IM phenotypes. Since *rs12248560* was not included in the exome capture, it had to be filled in using the array data.

### Gene-drug interaction analyses

We evaluated number of gene-drug interactions within the PMBB with medications with CPIC level A or B guidelines available. We extracted 16 loci (15 genes + the rs12777823 SNP) as listed in Table 3 that have gene-drug interactions. Among the ∼43,000 genotyped individuals, all individuals (100%) have at least one non-reference allele in the subset of loci we genotyped. PharmCAT annotation also identified that 98.9% of individuals are carriers of one or more PGx actionable phenotypes that would result in a pharmacotherapy modification. The mean number of genes per patient with an actionable phenotype was 4.1. A t-test comparing EUR (µ=4.088, n=30,008) and AFR (µ=4.282, n=11,156) actionable phenotype counts showed that AFR has a significantly higher mean count of phenotypes that correspond to treatment modification than EUR (p < 0.0001). The number of individuals stratified by each ancestry group for gene-phenotype combination are listed in Table 3. Next, we examined how many individuals with actionable alleles were prescribed drugs that would be impacted by those alleles as represented in Table 4. Among the top prescribed CPIC drugs where prescribed individuals have an actionable PGx allele are warfarin, clopidogrel, and omeprazole.

**Table 3:**
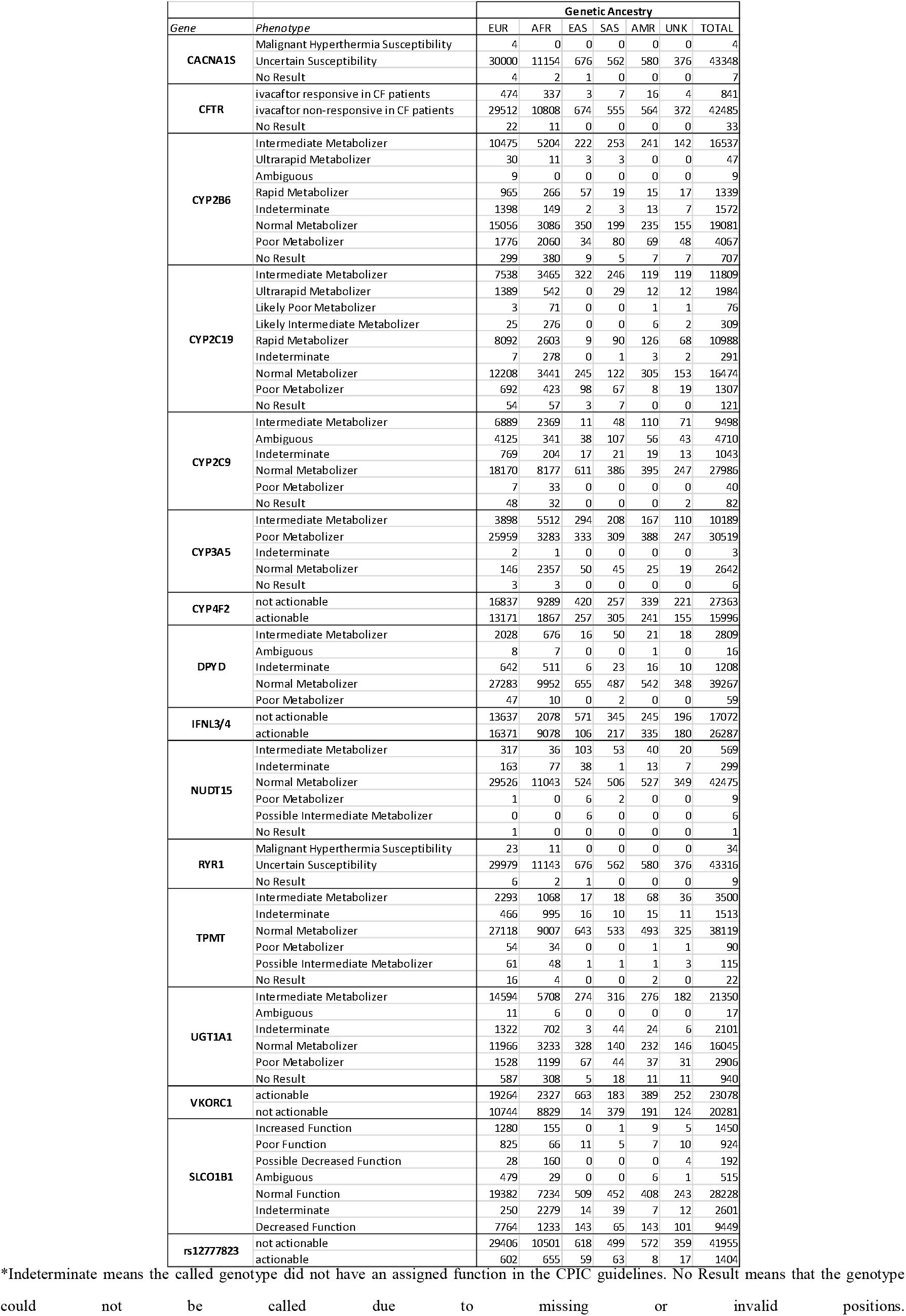
Number of participants in the PMBB with PGx phenotypes by ancestry based on PharmCAT annotation.

**Table 4:** Top 10 drugs by number of PMBB participants with PGx actionable alleles. Proportion of participants prescribed a medication also carrying an actionable allele for that drug. The count of prescribed drugs with actionable alleles (ACT), total prescribed drugs considered (TOT), and proportion of prescribed drugs with actionable alleles (PROP) are shown.

|  |  | Proportion of Total Prescribed with Actionable PGx |  |  |  |  |  |  |  |  |  |  |  |  |  |  |  |  |  |  |  |  |
| --- | --- | --- | --- | --- | --- | --- | --- | --- | --- | --- | --- | --- | --- | --- | --- | --- | --- | --- | --- | --- | --- | --- |
|  |  | EUR |  |  | AFR |  |  | EAS |  |  | SAS |  |  | AMR |  |  | UNKNOWN |  |  | TOTAL |  |  |
| Drug | Genes | ACT | TOT | PROP | ACT | TOT | PROP | ACT | TOT | PROP | ACT | TOT | PROP | ACT | TOT | PROP | ACT | TOT | PROP | ACT | TOT | PROP |
| *warfarin* | CYP2C9<br>CYP4F2<br>VKORC1<br>rs12777823 | 1904 | 2216 | 0.8592 | 416 | 864 | 0.4815 | 19 | 20 | 0.9500 | 15 | 18 | 0.8333 | 19 | 22 | 0.8636 | 23 | 26 | 0.8846 | 2396 | 3166 | 0.7568 |
| *clopidogrel* | CYP2C19 | 537 | 1988 | 0.2701 | 276 | 753 | 0.3665 | 10 | 18 | 0.5556 | 20 | 34 | 0.5882 | 3 | 25 | 0.1200 | 10 | 19 | 0.5263 | 856 | 2837 | 0.3017 |
| *tacrolimus* | CYP3A5 | 214 | 1494 | 0.1432 | 409 | 585 | 0.6991 | 15 | 23 | 0.6522 | 7 | 27 | 0.2593 | 17 | 39 | 0.4359 | 1 | 24 | 0.0417 | 663 | 2192 | 0.3025 |
| *simvastatin* | SLCO1B1 | 507 | 1806 | 0.2807 | 94 | 994 | 0.0946 | 11 | 28 | 0.3929 | 3 | 24 | 0.1250 | 3 | 27 | 0.1111 | 7 | 23 | 0.3043 | 625 | 2902 | 0.2154 |
| *amitriptyline* | CYP2C19<br>(CYP2D6 not used) | 219 | 511 | 0.4286 | 196 | 522 | 0.3755 | 0 | 7 | 0.0000 | 3 | 4 | 0.7500 | 2 | 15 | 0.1333 | 4 | 9 | 0.4444 | 424 | 1068 | 0.3970 |
| *omeprazole* | CYP2C19 | 237 | 4780 | 0.0496 | 149 | 3177 | 0.0469 | 0 | 79 | 0.0000 | 6 | 105 | 0.0571 | 3 | 121 | 0.0248 | 2 | 63 | 0.0317 | 397 | 8325 | 0.0477 |
| *ibuprofen* | CYP2C9 | 76 | 4178 | 0.0182 | 110 | 4137 | 0.0266 | 0 | 150 | 0.0000 | 0 | 116 | 0.0000 | 1 | 148 | 0.0068 | 3 | 113 | 0.0265 | 190 | 8842 | 0.0215 |
| *escitalopram* | CYP2C19 | 115 | 1619 | 0.0710 | 53 | 648 | 0.0818 | 4 | 20 | 0.2000 | 3 | 16 | 0.1875 | 1 | 33 | 0.0303 | 1 | 12 | 0.0833 | 177 | 2348 | 0.0754 |
| *voriconazole* | CYP2C19 | 140 | 247 | 0.5668 | 24 | 34 | 0.7059 | 1 | 1 | 1.0000 | 3 | 5 | 0.6000 | 3 | 5 | 0.6000 | 0 | 3 | 0.0000 | 171 | 295 | 0.5797 |
| *lansoprazole* | CYP2C19 | 93 | 1890 | 0.0492 | 58 | 1086 | 0.0534 | 0 | 26 | 0.0000 | 0 | 31 | 0.0000 | 1 | 34 | 0.0294 | 1 | 28 | 0.0357 | 153 | 3095 | 0.0494 |

Since our data show a higher frequency of *CYP2C19* reduced function alleles in East Asian ancestry individuals, we decided to see if this trend resulted in larger proportions of clopidogrel users of East Asian ancestry having actionable *CYP2C19* IM and PM phenotypes. We performed a Fisher’s exact test to compare these proportions of individuals of EUR and Asian ancestry (EAS & SAS) prescribed clopidogrel who had IM or PM phenotypes. In the EUR group there were 537 IMs and PMs (actionable for clopidogrel), and 1451 patients with nonactionable phenotypes. In the Asian ancestry group, there were 22 without and 30 with actionable phenotypes. We found a significantly larger proportion of Asian ancestry individuals taking clopidogrel are IMs and PMs than in EUR (p < 0.0001) with an odds ratio of 3.68, indicating that clopidogrel users of Asian ancestry are 3.68 times more likely than individuals of European ancestry to have actionable PGx alleles for clopidogrel.

Not all genes could be genotyped unambiguously for all patients due to missing alleles. PharmCAT provides output as several genotypes with the highest matching score. In many cases, the ambiguous genotypes (i.e. two or more genotypes) result in the same phenotype. In cases when they do not, the phenotype is ambiguous as well. For example, 10.9% of patients had an ambiguous *CYP2C9* phenotype, which likely resulted in an underestimate of patients with actionable *CYP2C9* phenotypes taking ibuprofen, the second most common PGx drug in Penn Medicine. A full breakdown of uncalled or ambiguous genotypes and phenotypes from PharmCAT is available in Supplementary Table 2.

### CYP2D6 analysis

Despite a lack of copy-number and structural variant information for *CYP2D6*, SNP-based genotyping may still provide some estimates into the burden of *CYP2D6* IM and PMs at the population level with the caveat that a number of no function alleles (defined by structural variants and the gene deletion *5) cannot be captured and samples including these are determined as IM or NM depending on the second allele. PharmCAT was able to call *CYP2D6* normal and reduced function phenotypes (NM/IM/PM) for 14,581 (33.6%) of the PMBB individuals; all other individuals were either Indeterminate due to the presence uncertain or unknown function alleles, or No Result for un-callable individuals (Supplementary Table 3). Of those called, 7,254 (16.7%) had reduced function phenotypes (IM/PM). If we calculate gene activity scores from diplotypes, only 14,460 (33.3%) are unambiguous since metabolizer phenotypes encompass a range of activity scores and some individuals with multiple possible genotypes can have multiple activity scores which all fall under the same metabolizer category (Supplementary Table 4). Using these phenotypes, we provided a lower-bound estimate of the number of patients receiving drugs for which they have an actionable *CYP2D6* IM/PM phenotype (Table 5). However, in the absence of copy number variation, we could not call RM or UM phenotypes, which are also actionable for many *CYP2D6*-influenced drugs. Despite these limitations, we were able to detect 792 patients prescribed codeine and 761 patients prescribed tramadol with an actionable IM/PM phenotype in *CYP2D6*. For ondansetron, our most prescribed PGx-affected drug, only the UM phenotype is considered actionable and therefore we could not assess this drug-gene interaction.

**Table 5.** Proportion of PMBB participants with actionable CYP2D6 IM/PM phenotypes from PharmCAT research-mode annotation for drugs they have been prescribed.

| *Drug* | Actionable | Total Prescribed | Proportion |
| --- | --- | --- | --- |
| *amitriptyline* | 160 | 1068 | 0.1498 |
| *atomoxetine* | 29 | 77 | 0.3766 |
| *codeine* | 792 | 5278 | 0.1501 |
| *nortriptyline* | 105 | 865 | 0.1214 |
| *paroxetine* | 9 | 489 | 0.0184 |
| *tamoxifen* | 42 | 304 | 0.1382 |
| *tramadol* | 761 | 5194 | 0.1465 |

## Discussion

The identification of many clinically actionable alleles and their interactions with medications has led investigators to critically think about implementing PGx testing of these alleles into clinical practice. Our study examined approximately 3.3 million individuals from Penn Medicine and identified that 21.3% of patients have been prescribed at least one CPIC guideline medication and 9.3% were prescribed two or more. This evidence suggests that the clinical testing of pharmacogenetic alleles could help in informing patient care at Penn Medicine and other large healthcare systems. Our analyses on ∼43,000 genotyped individuals from PMBB demonstrate that the frequency of participants with one or more clinically actionable alleles is very high. 100% of participants in our biobank population are carriers of at least one non-referent allele in a gene with a CPIC Level A or B guideline, out of which 98.9% of participants have a PGx phenotype that would lead to change in the prescription or dosage of at least one drug. More than 14% of patients were prescribed a drug for which they carry an actionable allele in our 8-year study period (2012-2020). These results suggest strong evidence for the benefits of integrating pharmacogenomic testing with current clinical practice for precision health care. Pharmacogenomic testing can be beneficial in reducing the number of drug adverse effects and nonresponse outcomes as well as minimize trial and error for dose specification. As more pharmacogenetic associations are discovered for more drugs, and as existing PGx-affected medications continue to grow in usage, the utility and clinical impact of pharmacogenomic infrastructure will only grow. At scale, these interventions can be a powerful and cost-effective method for improving patient care.

In this study, we aimed to observe the landscape of prescription trends of drugs that have a pharmacological impact in the Penn Medicine EHR to make necessary decisions about where and how to implement pharmacogenomics. Similar findings have also been reported by several other previous studies(10,11,14,18) but to our knowledge, our study is unique in analyzing pharmacogenomic alleles among already genotyped individuals across multiple ancestry groups for identifying individuals that could benefit from pharmacotherapy. We evaluated the potential impact of PGx testing by retrospective analyses in our PMBB population and compared our findings to similar studies in one or multiple health systems. We identified different challenges in collecting and mining data from the EHR to observe these trends. Our analyses aimed at identifying individuals who could benefit from pharmacogenomic testing while highlighting essential considerations for clinicians and researchers for implementing pharmacogenomics in clinical practice. Genotyping individuals with a one-time panel of PGx genes would impact medication prescribing over the course of care. Among the most prescribed medication classes affected by PGx alleles include pain medications, anti-emetics, and proton pump inhibitors. These medications are often prescribed in the post-operative or acute care setting and represent a high-risk patient population that will benefit from implementation of pre-emptive PGx panel testing.(19) In one retrospective analysis in patients who underwent panel based PGx testing, the presence of a gene-drug interaction for CPIC guideline medications was associated with an increased risk of 90-day hospital readmission by more than 40%.(20) An ongoing prospective study will determine whether PGx panel testing is feasible in the acute care setting.**(21)**

Our study examined the evidence based CPIC guidelines and applied them in a clinical setting to quantify their burden on actual patients. Based on the widespread prevalence of PGx alleles and PGx-affected medication, there would be substantial benefit to preemptive, universal PGx testing in both patient outcomes and healthcare costs. Warfarin is a prime example for a drug in widespread use where genetic information may inform dosing in a large proportion of patients. Prior knowledge of warfarin responsiveness can reduce trial and error in dose titration towards a safe and effective dose. Three randomized controlled trials of PGx guided warfarin dosing have been performed and one found PGx testing to increase the time in therapeutic range(22) and one of the studies showed an improvement in the composite clinical outcome of major bleeding, supratherapeutic International Normalized Ratio (INR), venous thromboembolism, or death(23). While the use of reactive PGx testing may not be clinical feasible, the use of preemptive testing for PGx genes to guide cardiovascular medications including warfarin was found to be cost-effective.(24) In addition to dose-modifying effects, pharmacogenomic analyses have identified many gene-drug interactions such as clopidogrel where patient carrying an alternate allele is entirely indicated against taking a drug due to high risk of nonresponse leading to adverse outcomes. Nonresponse to clopidogrel treatment resulting in stroke or myocardial infarction is a severe outcome for patients that leads to dramatic economic costs but is preventable. Information on an individual patient’s drug metabolizer status can aid clinicians in making informed decisions regarding which drug regimen should be selected for patients based on their genetic information. Owing to the ancestry diversity of the PMBB, we were able to demonstrate that the burden of PGx alleles disproportionately affects certain ancestry groups. We report notable differences in phenotype frequency for many of the studied genes in each ancestry group. In aggregate, European ancestry individuals have significantly lower average counts of actionable alleles than individuals of African ancestry (p<0.0001), as well as compared to all non-European ancestry combined. Furthermore, patients of Asian ancestry taking clopidogrel therapy have greatly increased rates of treatment modifying *CYP2C19* reduced metabolizer phenotypes than individuals of European ancestry, quantifying a burden of PGx in actual patients. IM and PM phenotypes may result in increased risk of major adverse cardiovascular events in clopidogrel users, and therefore this demonstrates an unequal burden of PGx-related harms in clopidogrel patients of Asian ancestry. We found similar trends of differential PGx burden across ancestry groups for other widely used drugs including tacrolimus and voriconazole. Despite the observation of ancestry-specific patterns in PGx, we strongly discourage the use of ancestry as a proxy for PGx testing. In the case of clopidogrel, although a majority of clopidogrel-prescribed patients of Asian ancestry have actionable PGx alleles in *CYP2C19*, 42.3% still do not. Furthermore, in clopidogrel-prescribed patients of European ancestry, a still-notable 27.0% of patients have actionable *CYP2C19* alleles. These findings demonstrate how inaction towards the implementation of pharmacogenetic testing harms all patients, but disproportionately harms patients of non-European ancestry.

Although our study highlighted a large proportion of pharmacogenomic alleles with CPIC guidance in the PMBB population, several limitations should be noted. We identified variations based on an integrated call set of genotype chip and whole exome data in VCF format, which does not capture PGx-relevant copy number or structural variations (in *CYP2D6*) and is also not well suited to calling *HLA* polymorphisms. As a result, we used presumptive *CYP2D6* calls from PharmCAT which must be interpreted carefully. Another limitation is that although we were able to collect data on the prescribed medications during encounters at Penn Medicine, we are unable to confirm if the prescription is new or was prescribed at another clinical care setting. Thus, identification of patients with adverse reactions to drugs with CPIC Level A or B guidelines from the EHR has also been challenging. To address this, future work could involve developing algorithms for identifying previously administered drugs by mining through patient notes and EHR fields, and mining through longitudinal EHR data, allergy fields, and notes for adverse reactions and nonresponse to drugs. This would allow us to further quantify the burden of PGx as a measure of actual health outcomes. It should also be noted that although the integrated call set allowed us to genotype and average of 66% of PGx sites included in CPIC Level A or B guidelines, the missing sites in some cases limit our ability to call phenotypes resulted in some patients having missing or ambiguous calls for certain genes. Targeted PGx panels or whole-genome sequencing could largely avoid this issue, but targeted panels would eventually become obsolete as new alleles are discovered. Our study is unique in that it specifically explores how ancestry-specific patterns in PGx allele frequencies are reflected in patients receiving drugs for which they have a gene-drug interaction. Although we have more than 500 patients in every studied ancestry group which is enough to confidently demonstrate differences in phenotype frequencies, the relatively small proportion of patients who have been prescribed each drug limits our power to draw conclusions about ancestry-specific drug-gene interactions. As more patients are enrolled and genotyped by PMBB, and other data becomes available in resources like All of Us (citation of the cohort program), we will gain more power to disentangle these ancestry-specific associations, which carry major implications for the effect of PGx across different ancestry groups. It is important to consider relevant PGx alleles in all ancestry groups as more health systems consider clinical implementation of PGx; broad, diverse inclusion of multiple ancestry groups in clinical implementation has the potential to reduce health disparities. Lastly, the CPIC guidelines are easily accessible, but implementation of these guidelines poses several challenges in a clinical setting. One important challenge is the ability to return timely genotyping results prior to prescription of the drug. Based on our findings, it seems appropriate to perform genetic testing on patients preemptively and universally before they are prescribed a relevant PGx-informed drug. However, this poses many challenges which include but are not limited to cost effectiveness, insurance coverage, future discovery of new actionable genes and alleles, integration of real time clinical decision support, availability of resources, patient-prescriber education, and review of genetic reports among others.

## Conclusions

With the increasing availability of clinical decision support tools like PharmCAT, it will soon be possible to add pharmacogenetic information to EHR systems such as Epic. With proper education and resources, along with increased adoption of PGx testing, healthcare professionals will be able to utilize this information to make genetics-informed drug dosing and prescription. Addressing these barriers will be imperative before widespread adoption of clinical PGx can be achieved. Implementation of PGx into clinical workflows has the potential to affect clinical care for a very large proportion of the health care population and is a significant contributor to fully realizing the practice of precision medicine.

## Data Availability

All data produced in the present work are contained in the manuscript

## List of Abbreviations

(PGx): Pharmacogenomics
(CPIC): The Clinical Pharmacogenetics Implementation Consortium
(EHRs): Electronic Health Records
(PMBB): The Penn Medicine BioBank
(SNP): Single Nucleotide Polymorphism
(WES): Whole Exome Sequencing
(PharmCAT): Pharmacogenomics Clinical Annotation Tool
(VCF): Variant call format
(PDS): Penn Data Store
(PM): Poor Metabolizer
(IM): Intermediate Metabolizer
(NM): Normal Metabolizer
(RM): Rapid Metabolizers
(UM): Ultrarapid Metabolizer
(EUR): European ancestry
(AFR): African ancestry
(EAS): East Asian ancestry
(AMR): Admixed American ancestry
(SAS): South Asian ancestry
(UNK): Unknown ancestry
(NSAIDs): Non-Steroidal Anti-Inflammatory Drugs

## Acknowledgements

This work was supported by the Penn Center for Precision Medicine Accelerator Fund. Additional funding provided by K23HL143161 for Dr. Sony Tuteja and PharmCAT grant: NHGRI U24HG010862

## Competing Interests

MDR was on the Scientific Advisory Board for Cipherome until June 2022. All other authors declare no competing interests.

**Supplementary Figure 1:**
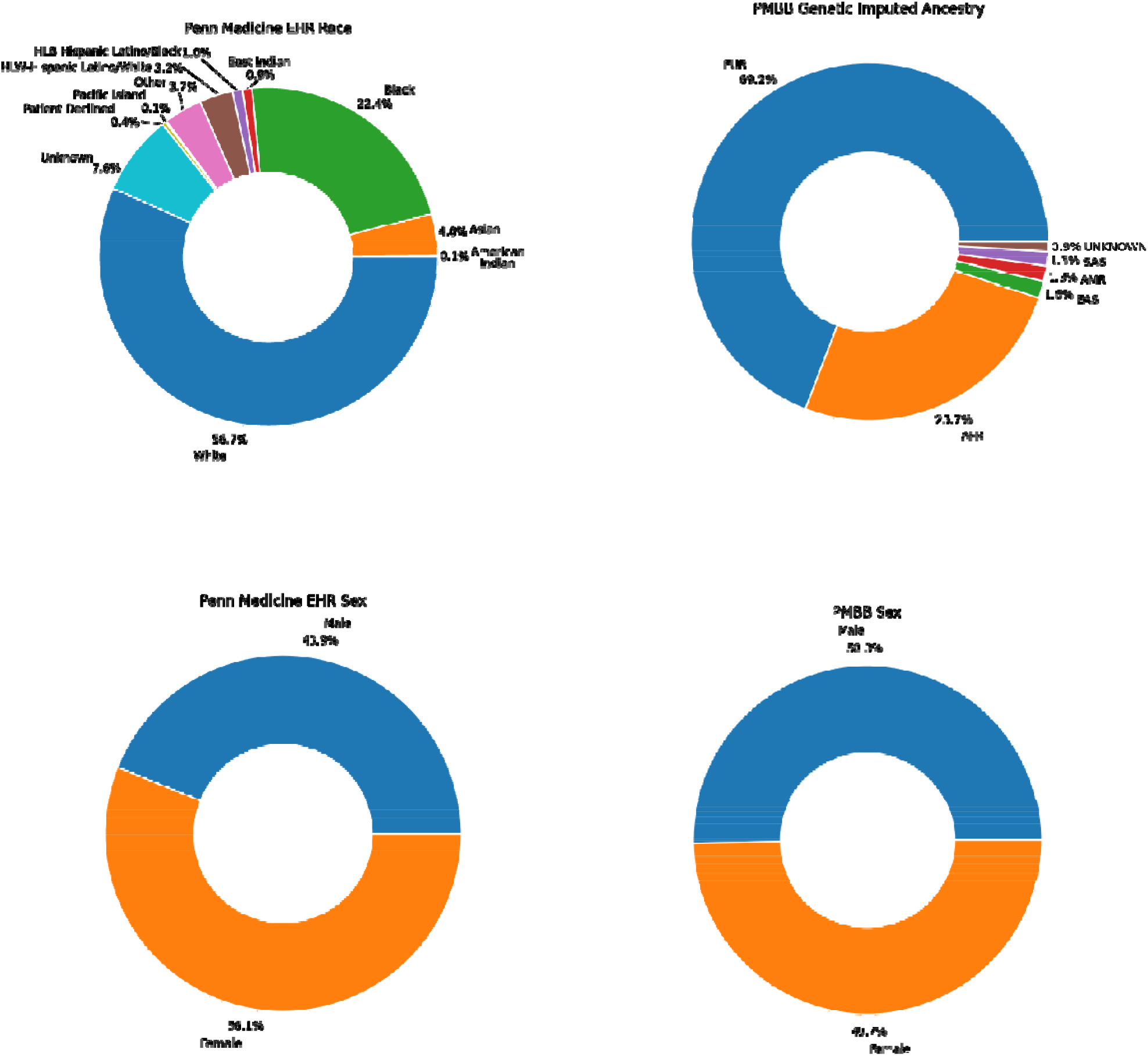
Demographic distribution on Penn Medicine EHR on left and Penn Medicine Biobank on the right.

**Supplementary Table 1:** Drugs queried for the study See *cpic_actionable*.*xlsx*

| **Medication** | **Indication** | **CPIC classification** |
| --- | --- | --- |
| Abacavir | HIV | A |
| Allopurinol | Gout | A |
| Amitriptyline | Depression | A |
| Atazanavir | HIV | A |
| Atomoxetine | ADHD | A |
| Azathioprine | Immunosuppression | A |
| Capecitabine | Cancer | A |
| Carbamazepine | Seizures | A |
| Celecoxib | Pain | A |
| Citalopram | Depression | A |
| Clomipramine | Depression | B |
| Clopidogrel | Myocardial infarction/stroke | A |
| Codeine | Pain | A |
| Desipramine | Depression | B |
| Dexlansoprazole | Acid reflux | B |
| Doxepin | Depression | B |
| Efavirenz | HIV | A |
| Escitalopram | Depression | A |
| Fluorouracil | Cancer | A |
| Flurbiprofen | Pain | A |
| Fluvoxamine | Depression | B |
| Fosphenytoin | Seizures | A |
| Hydrocodone | Pain | B |
| Ibuprofen | Pain | A |
| Imipramine | Depression | B |
| Ivacaftor | Cystic Fibrosis | A |
| Lansoprazole | Acid reflux | A |
| Meloxicam | Pain | A |
| Mercaptopurine | Immunosuppression | A |
| Nortriptyline | Depression | A |
| Omeprazole | Acid reflux | A |
| Ondansetron | Nausea/vomiting | A |
| Oxcarbazepine | Seizures | A |
| Pantoprazole | Acid reflux | A |
| Paroxetine | Depression | A |
| Peginterferon alfa-2a | Hepatitis C | A |
| Peginterferon alfa-2b | Hepatitis C | A |
| Phenytoin | Seizures | A |
| Piroxicam | Pain | A |
| Rasburicase | Hyperuricemia | A |
| Sertraline | Depression | B |
| Simvastatin | Hypercholesterolemia | A |
| Succinylcholine | Paralytic |  |
| Tacrolimus | Immunosuppression | A |
| Tamoxifen | Cancer | A |
| Thioguanine | Cancer | A |
| Tramadol | Pain | A |
| Voriconazole | Fungal infection | A |
| Warfarin | Thromboembolic disorders | A |

**Supplementary Table 2:** Numbers of samples with ambiguous and uncalled genotypes and phenotypes from PharmCAT

| *Gene/SNP* | # Not Called* | Proportion Not Called | # Ambiguous Genotype** | # Ambiguous Phenotype** | Proportion Ambiguous Genotype | Proportion Ambiguous Phenotype |
| --- | --- | --- | --- | --- | --- | --- |
| **CACNA1S** | 7 | 0.0002 | 0 | 0 | 0 | 0 |
| **CFTR** | 33 | 0.0008 | 0 | 0 | 0 | 0 |
| **CYP2B6** | 707 | 0.0163 | 13221 | 9 | 0.3049 | 0.0002 |
| **CYP2C19** | 121 | 0.0028 | 226 | 0 | 0.0052 | 0 |
| **CYP2D6** | 13536 | 0.3122 | 5984 | 2701 | 0.138 | 0.0623 |
| **rs12777823** | 1701 | 0.0392 | 0 | 0 | 0 | 0 |
| **CYP2C9** | 82 | 0.0019 | 4799 | 4710 | 0.1107 | 0.1086 |
| **CYP3A5** | 6 | 0.0001 | 0 | 0 | 0 | 0 |
| **CYP4F2** | 2679 | 0.0618 | 0 | 0 | 0 | 0 |
| **DPYD** | 16 | 0.0004 | 0 | 0 | 0 | 0 |
| **IFNL3** | 2265 | 0.0522 | 0 | 0 | 0 | 0 |
| **NUDT15** | 1 | 0 | 0 | 0 | 0 | 0 |
| **RYR1** | 9 | 0.0002 | 0 | 0 | 0 | 0 |
| **TPMT** | 22 | 0.0005 | 1 | 0 | 0 | 0 |
| **UGT1A1** | 940 | 0.0217 | 50 | 17 | 0.0012 | 0.0004 |
| **VKORC1** | 70 | 0.0016 | 0 | 0 | 0 | 0 |
| **SLCO1B1** | 0 | 0 | 917 | 515 | 0.0211 | 0.0119 |

\*PharmCAT may fail to call samples when variants are missing from the input samples, either due to lack of coverage from the genotyping method or due to filtering during quality control. Furthermore, it can fail to call when the variants cannot be matched to an exact genotype in the allele definitions.
\*\*Ambiguous genotypes are samples for which PharmCAT returned two or more genotypes with the same score based on matching the variant input to the allele definitions. An allele can be defined by one or more variant(s) and a variant can be present in several alleles. In case of unphased data, there are instances for which a variant input from the VCF file might match more than one genotype given the overlap in allele definitions. If PGx relevant positions are missing in the input data, those positions are ignored in the allele definitions. If that covers all variants in an allele definition, the allele cannot be considered in the matching process. If it only affects part of the variants in an allele definition, the allele is included in the matching process based on the remaining positions. Given the above-described overlap in allele definitions, missing positions can lead to definitions not being distinct from each other and the PharmCAT matcher provides all possible combinations with the highest score. Subsequently the PharmCAT Phenotyper assigns each genotype a corresponding phenotype based on the information stored in the CPIC database. In cases with two or more genotypes (ambiguous genotypes) the corresponding phenotypes were compared and if the phenotypes differ the sample was labeled as ambiguous phenotype. The above table lists the numbers of samples with ambiguous geno- and phenotypes. These samples were excluded from the analyses presented in the manuscript.

**Supplementary Table 3:** *CYP2D6* phenotype counts from PharmCAT research mode (without CNV and structural variants)

| *CYP2D6* Phenotype | Count |
| --- | --- |
| Normal Metabolizer | 7327 |
| Intermediate Metabolizer | 6330 |
| Poor Metabolizer | 924 |
| Ambiguous | 2701 |
| Indeterminate | 12541 |
| No Result | 13536 |

**Supplementary Table 4:**
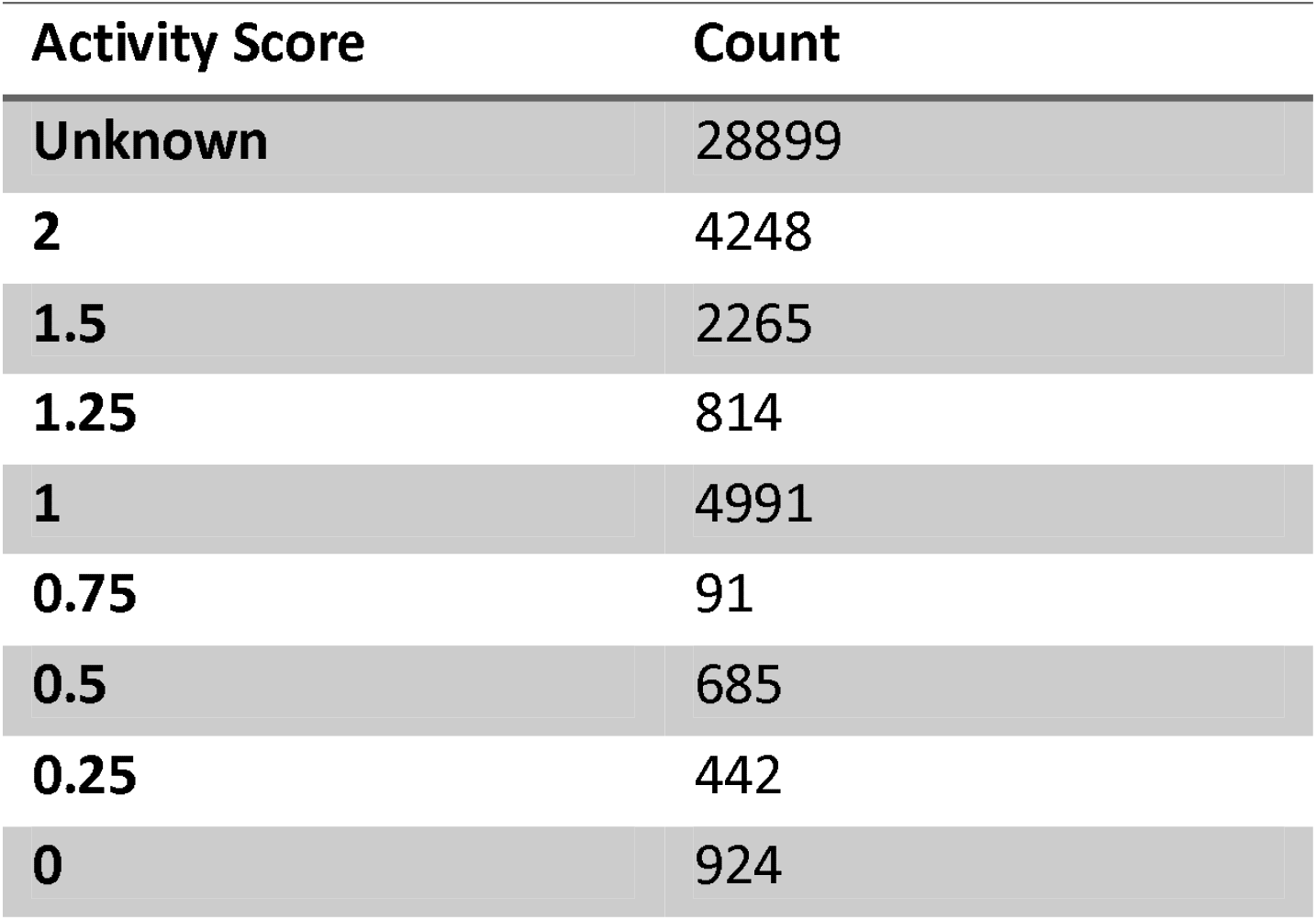
*CYP2D6* activity score counts from PharmCAT research mode (without CNV and structural variants)

| Activity Score | Count |
| --- | --- |
| Unknown | 28899 |
| 2 | 4248 |
| 1.5 | 2265 |
| 1.25 | 814 |
| 1 | 4991 |
| 0.75 | 91 |
| 0.5 | 685 |
| 0.25 | 442 |
| 0 | 924 |

## Notes

### Competing Interest Statement

MDR was on the Scientific Advisory Board for Cipherome until June 2022. All other authors declared no competing interests.

### Author Declarations

The research study complies with the Declaration of Helsinki and the research was approved by the University of Pennsylvania Institutional Review Board. Participants provided informed consent prior to enrollment into the Penn Medicine Biobank and genetic analysis (IRB #813913, IRB #817977, IRB #808346). The IRB approved medical records extraction for the prescription medication data under a waiver of informed consent (IRB #826757).

